# Human CART22.19 Therapy in Refractory Pediatric B-ALL: Insights from a Named-Patient Cohort

**DOI:** 10.1101/2025.09.09.25335341

**Authors:** Anna-Sophia Mast, Peter Lang, Patrick Schlegel, Friso G Calkoen, Daniel Atar, Sophia Scheuermann, Sylvia Klein, Christiane Braun, Florian Schinle, Marina Schmidt, Luca Hensen, Martin Ebinger, Michaela Döring, Jürgen Schäfer, Johannes H Schulte, Bader Alahmari, Peirong Hu, Dina Schneider, Rimas Orentas, Rupert Handgretinger, Christian M. Seitz

**Affiliations:** Department of General Pediatrics, Hematology and Oncology, University Children’s Hospital Tuebingen, Germany; Department of Pediatric Hematology, Oncology and Stem Cell Transplantation, University of Regensburg, Germany; Leibniz Institute for Immunotherapy, Regensburg, Germany; Princess Máxima Center for Pediatric Oncology, Utrecht, the Netherlands; Hopp-Children’s Cancer Center Heidelberg (KiTZ), Heidelberg, Germany; B310 Clinical Cooperation Unit Pediatric Oncology, German Cancer Research Center (DKFZ), Heidelberg, Germany; Department of Pediatric Oncology, Hematology, and Immunology, Heidelberg University Hospital, Heidelberg, Germany; German Cancer Research Consortium (DKTK), Partner Site Tuebingen, German Cancer Research Center (DKFZ), Heidelberg, Germany; Excellence cluster iFIT (EXC 2180) “Image-Guided and Functionally Instructed Tumor Therapies” Tuebingen, Germany; Department of Hematology and Oncology, University Hospital Tübingen, Tuebingen, Germany; Department of Diagnostic and Interventional Radiology, University Hospital Tuebingen, Tuebingen, Germany; Department of Oncology, Ministry of the National Guard-Health Affairs, Riyadh, Saudi Arabia; King Abdullah International Medical Research Center, Riyadh, Saudi Arabia; Lentigen, a Miltenyi Biotec Company, Gaithersburg, MD, USA; Research and Development Immunotherapy, Miltenyi Biotec, Bergisch Gladbach, Germany

## Abstract

**Background:** CD19-directed chimeric antigen receptor (CAR) T-cell therapies have transformed the treatment landscape for pediatric B-cell acute lymphoblastic leukemia (B-ALL), yet relapse driven by antigen escape remains a major limitation. Dual-targeting CAR approaches recognizing CD19 and CD22 have shown promising clinical activity. However, sustained remissions are limited by insufficient CAR T-cell persistence.

**Methods:** CAR22.19, a fully human tandem CD19/CD22 CAR, was developed and clinically applied within a named-patient program in nine heavily pretreated pediatric patients with relapsed/refractory B-ALL. Treatment indications were CD19-negative blast population (n=5), relapse after CD19 CAR T (n=3) and/or restricted access to approved CAR T-cell products (n=3). Autologous and donor-derived CAR22.19 T-cells (CART22.19) were manufactured using a GMP-compliant, semi-automated fresh in fresh out process. Safety and efficacy were assessed through standardized clinical monitoring, measurable residual disease analysis, and CAR T-cell kinetics.

**Results:** Preclinical validation demonstrated antigen-specific cytotoxicity and dual-antigen activity. Clinically, CART22.19 were well tolerated, with no treatment-related deaths and no grade ≥3 neurotoxicity, while grade ≥3 cytokine release syndrome occurred in 38.5% (5/13) of infusions and resolved with standard interventions. An initial complete molecular remission was achieved in 78% (7/9) of patients, with a 12-month overall survival rate of 53.3% (95% CI, 17.7-79.6%). Sustained treatment response in CD19⁻CD22⁺ cases underscore the functional contribution of the CD22-targeting domain. In contrast, all patient’s refractory to prior CD19 CAR T-cell therapies relapsed early with retained CD19⁺CD22⁺ expression. Limited *in vivo* persistence was found to be a key mechanism of treatment failure. Notably, durable remission and sustained functional persistence of CART22.19 was achieved in one patient refractory to autologous CART22.19 following infusion of donor-derived CART22.19 after reduced-intensity conditioning (RIC) allogeneic hematopoietic stem cell transplantation (alloHSCT) in non-remission.

**Conclusions:** CART22.19 therapy demonstrated a favorable safety profile and promising clinical activity in a high-risk pediatric population, with dual targeting enabling disease control in CD19-negative disease. However, limited CAR T-cell persistence remains a major obstacle to sustained remission. Our findings support further clinical development of CART22.19 and highlight the potential of donor-derived CAR T-cells following RIC alloHSCT as a novel therapeutic strategy to enhance persistence and improve outcomes in heavily pretreated pediatric patients.

## Introduction

The advent of CD19-targeted CAR T-cells (CAR19) has revolutionized the therapy of pediatric acute lymphoblastic leukemia (ALL) (1–5). While initial complete remission rates are high (70-90%), the interpretation of long-term event-free survival (EFS) remains challenging due to heterogeneity in post-remission treatment strategies. Specifically, a variable proportion of patients (13-88%) undergo consolidative allogeneic hematopoietic stem cell transplantation (alloHSCT), which complicates outcome comparisons across studies (2, 5, 6).

Relapse following CAR19 therapy generally manifests in two distinct patterns: CD19-negative relapse, often attributed to effective elimination of CD19-positive cells by potent CAR T-cell products, and CD19-positive relapse, which is associated with limited CAR T-cell functionality or persistence (3, 4, 7–14). Antigen escape, characterized by the loss or modulation of the targeted surface antigen, poses a significant challenge in both CD19- and CD22-targeted CAR T-cell therapies (3, 15, 16).

To counteract antigen-negative escape, dual-targeting CAR T-cell approaches have been developed, particularly combining CD19 and CD22. These can be implemented through several strategies: (1) co-administration of two monospecific CAR T-cell products, (2) co-transduction of T-cells with two separate vectors, (3) adapter CAR platforms that enable modular antigen recognition, (4) use of bicistronic vectors encoding both CARs, or (5) tandem CAR constructs incorporating two antigen-binding domains into a single receptor (17–23).

While co-administration and co-transduction strategies facilitate rapid translation using validated components, they yield heterogeneous cell populations and increase manufacturing complexity. In contrast, bicistronic and tandem formats enable the production of uniform, dual-specific CAR T-cell products in which every transduced cell targets both antigens, offering potential manufacturing and cost advantages.

Despite promising safety data, dual CD19/CD22-directed CAR T-cell therapies have yet to demonstrate consistent superiority over monospecific CAR19 with respect to long-term outcomes. Reported 1-year EFS rates range widely from 32% to 83% (17–21, 24). Limited CAR T-cell persistence is frequently cited as a major contributor to relapse, although the mechanisms underlying CD19-positive relapse remain incompletely understood (17–19, 21, 25). Contributing factors may include prior cytotoxic treatments, T-cell exhaustion, or immune-mediated rejection.

We describe the development and clinical application of CART22.19, a novel fully human tandem CAR targeting CD19 and CD22. Nine pediatric patients with relapsed or refractory B-ALL received CART22.19 therapy within a named-patient program. We provide an overview of clinical outcomes in the cohort, followed by an in-depth case description of a patient who achieved long-term remission after receiving donor-derived CART22.19 following reduced-intensity conditioning (RIC) alloHSCT in non-remission.

## Methods

### Design of the CAR22.19 Construct

The fully human CAR22.19 construct consists of a tandem targeting domain, comprising an anti-CD22 single-chain variable fragment (scFv; clone 16p17) fused in-frame to an anti-CD19 scFv (clone M19217-1). This tandem scFv is linked to a CD8α hinge and transmembrane region, followed by a 4-1BB costimulatory domain and a CD3ζ signaling domain. A schematic representation of the CAR22.19 structure is provided in **Figure 1A**.

**Figure 1:**
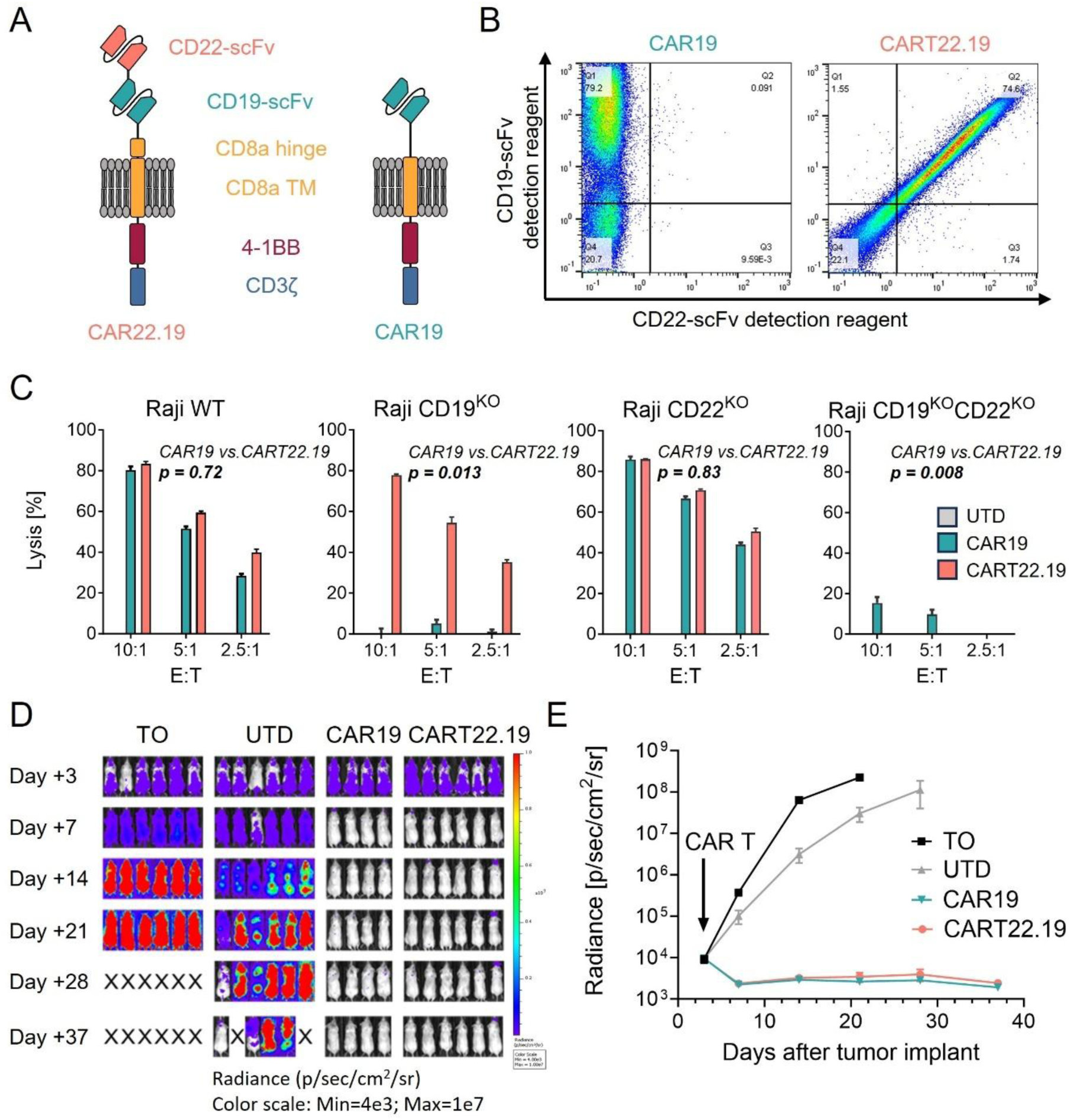
Design and Pre-Clinical Evaluation of CAR22.19. **A** Schematic depiction of the fully human CAR22.19 and a conventional CAR19. CAR22.19 comprises an anti-CD22 scFv linked in-frame to an anti-CD19 scFv. **B** Surface expression of CAR constructs was assessed by flow cytometry using binder-specific detection reagents for the CD19- and CD22-targeting scFv domains. **C** CART22.19, CAR19, or UTD were co-cultured with target cells at indicated E:T ratios for 16 hours. Target cell lysis was quantified by luminometry to determine the percentage of cytotoxicity. Targets were Raji WT (CD19⁺CD22⁺), Raji CD19^KO^ (CD19⁻CD22⁺), Raji CD22^KO^ (CD19⁺CD22⁻), and Raji CD19^KO^CD22^KO^ (CD19⁻CD22⁻). Values represent the mean ± SD of three technical replicates. Statistical significance was assessed using unpaired t-tests. **D-E** *In vivo* evaluation of CART22.19 in a NALM-6 xenograft model. NSG mice received 1×10⁶ firefly luciferase-expressing NALM-6 cells intravenously on day 0. On day 3, 1×10^7^ effector T-cells (CART22.19, CAR19 or UTD) were administered intravenously. Tumor burden was monitored by BLI, and mice were euthanized upon meeting predefined endpoint criteria. **D** Representative BLI images at indicated time points. **E** Quantification of BLI radiance over time. Data represent mean ± SEM; n=4-6 mice per group. Abbreviations: BLI: bioluminescence imaging; CAR: chimeric antigen receptor; CAR19: CD19-directed CAR; CAR22.19: fully human tandem CAR with anti-CD22 and anti-CD19 scFvs; CART22.19: CAR22.19 T-cells; E:T: effector-to-target; KO: knockout; NSG: NOD scid gamma; scFv: single-chain variable fragment; SD: standard deviation; SEM: standard error of the mean; TO: tumor only control; TM: transmembrane domain; t-test: unpaired Student’s t-test; UTD: untransduced activated T-cells; WT: wild type.

### Clinical Scale Manufacturing of CART22.19

The investigational product CART22.19 was manufactured from autologous or allogeneic T cells obtained by leukapheresis from patients or their HSCT donors, processed fresh without cryopreservation, and genetically modified to express the CAR22.19 construct. Manufacturing was performed under cleanroom conditions at the GMP-certified facility of the University Hospital Tübingen (UKT), following a standardized 12-day T-cell culture and transduction protocol on the CliniMACS Prodigy™ platform (Miltenyi Biotec), as previously described (26). A GMP-grade, second-generation lentiviral vector encoding the CAR22.19 construct under the control of the human EF-1α promoter was kindly provided by Miltenyi Biotec.

All CAR T-cell products, administered fresh without cryopreservation, underwent comprehensive quality control testing, including assessments of sterility, cell viability, transduction efficiency, and other predefined release criteria. Summary data from in-process monitoring and final product testing are provided in **Supplementary Table S1**. Additional information on lentiviral vector production and preclinical validation (*in vitro* and *in vivo*) is included in the Supplementary Methods.

### Patient Cohort

This report summarizes outcomes from individually authorized named-patient treatments administered under the hospital exemption provision for advanced therapy medicinal products (ATMP) (Article 28(2) of Regulation (EC) No 1394/2007). Informed consent was obtained from all patients or their legal guardians following detailed counseling by the treating physicians. Ethics committee approval was not required under the German Hospital Exemption pathway. All patients had relapsed or refractory disease with no remaining standard treatment options and met at least one of the following criteria: relapse following prior therapy with an approved CAR19 product, presence of CD19-negative leukemic blast populations, or lack of access to a licensed CAR19 therapy. Treatment with CART22.19 was recommended by the interdisciplinary Cell Therapy Board at the University Hospital Tübingen (UKT).

Autologous CART22.19 infusions were administered to patients without a history of HSCT or lacking access to the original donor (n = 7), whereas donor-derived allogeneic infusions were given to patients with prior allogeneic HSCT and an available original donor (n = 3). Upon relapse, two patients received two additional CART22.19 infusions from cryopreserved products as approved by the Cell Therapy Board, resulting in a total of 13 infusions across 9 patients.

### Reduced-Intensity Conditioning

Reduced-intensity conditioning (RIC) in patient P1 consisted of fludarabine (120 mg/m² body surface area (BSA)), melphalan (140 mg/m² BSA), and anti-thymocyte globulin (ATG; 30 mg/kg body weight (BW)), followed by allogeneic HSCT from a 9/10 HLA-matched unrelated donor. A TCRαβ/CD19-depleted graft was used, and no additional systemic immunosuppression was administered.

### CAR Administration

Lymphodepleting chemotherapy was initiated between day −12 and day −4 and consisted of fludarabine (Flu) at 25 mg/m² BSA once daily for three consecutive days, combined with a single dose of cyclophosphamide (Cy) at 900 mg/m² BSA. In select cases, the regimen was adjusted at the discretion of senior physicians based on individual risk factors and anticipated toxicity. CART22.19 was infused on day 0 at a mean dose of 2.9 × 10⁶ cells/kg BW (range, 0.3-6.0 × 10⁶ cells/kg BW).

### Therapy Response and Safety Assessment

Treatment response was evaluated by bone marrow minimal residual disease (MRD) analysis using real-time quantitative PCR (RQ-PCR) or multiparameter flow cytometry, and by imaging studies in cases with extramedullary involvement. Complete remission (CR) was defined as MRD negativity in the bone marrow and the absence of extramedullary disease. Treatment-related toxicities, including cytokine release syndrome (CRS) and immune effector cell-associated neurotoxicity syndrome (ICANS), were graded according to the criteria established by the American Society for Transplantation and Cellular Therapy (ASTCT) and the Common Terminology Criteria for Adverse Events (CTCAE), version 5.0 (27, 28). Laboratory parameters were monitored routinely during treatment and follow-up to assess toxicity and treatment response. Detailed methodologies are described in the Supplementary Methods.

### Outcomes

EFS was defined as the time from CART22.19 infusion to the first documented event: MRD reappearance, clinical relapse, or death from any cause. In patients who received more than one CART22.19 infusion, EFS was calculated from the date of the most recent administration. Patients undergoing subsequent HSCT were not censored. Overall survival (OS) was measured from the date of the first CART22.19 infusion to death from any cause. Duration of remission was defined as the time from achievement of complete remission to either relapse or death attributable to ALL. For patients who received more than one CART22.19 infusion, remission duration was calculated from the most recent infusion.

### Data Analysis

Objective response rates as well as EFS and OS rates were reported with two-sided 95% confidence intervals (CIs), calculated using the Clopper-Pearson method. Survival curves were estimated using the Kaplan-Meier method. All subgroup analyses were conducted post hoc and reported descriptively due to the small sample size and non-interventional nature of the dataset. No formal hypothesis testing was performed. Statistical significance was defined as p < 0.05. Analyses were performed using GraphPad Prism (version 9.4.1), and flow cytometry data were processed with FlowJo (version 10.8).

## Results

### Pre-Clinical Evaluation of CART22.19

To overcome antigen escape through CD19 loss and to additionally reduce immunogenicity associated with murine-derived single-chain variable fragments (scFvs), a fully human tandem CAR22.19 was developed (**Fig. 1A**). Transduction of T-cells with the CAR22.19 construct resulted in robust surface expression, with both binding domains assessable for target binding (**Fig. 1B**). Functional characterization demonstrated antigen-specific cytotoxicity and cytokine secretion (IL-2, IFN-γ) in response to CD19⁺CD22⁺ Raji cells as well as CD19 or CD22 single-knockout variants, but not to double-knockout cells lacking both target antigens (**Fig. 1C**; **Suppl. Fig. S1A, B**). In contrast to monospecific CAR19 T-cells, responsive only in the presence of CD19, CART22.19 are activated via either antigen, CD19 or CD22.

*In vivo*, CART22.19 induced rapid and sustained tumor clearance, comparable to CAR19 T-cells (**Fig. 1D, E**) and persisted with a predominant central memory phenotype (**Suppl. Fig. S1C, D**). These results strongly supported further clinical development of CART22.19 and prompted the initiation of a named-patient program.

### Patient Characteristics

Based on the promising preclinical results, nine pediatric patients with relapsed or refractory B-ALL were treated with CART22.19 within a named-patient program between November 2019 and June 2024. All patients had exhausted standard therapeutic options and were ineligible for approved commercial CAR T-cell therapies. Five patients presented with CD19-negative leukemic blast populations, three had relapsed after previous CAR19 therapy, and in three cases, regulatory constraints precluded access to commercial products.

CART22.19 were manufactured using a semi-automated process on the CliniMACS Prodigy platform, achieving a median transduction efficiency of 53% (range, 48.3-70.8%) (**Suppl. Table S1**). Thirteen infusions were administered across the nine patients, with two individuals receiving three separate infusions due to persistent or recurrent disease. Seven patients received autologous CAR T-cell products, while three were treated with donor-derived CART22.19. The most common administered dose was 3 × 10⁶ CART22.19/kg BW, used in eight of the thirteen infusions, with an overall dosing range of 0.3 × 10⁶ to 6 × 10⁶ CART22.19/kg BW.

Baseline patient characteristics are detailed in **Table 1**. The median age at initial diagnosis was 10.2 years (range, 0.2-16.6 years). All patients were heavily pretreated, with a median of four prior therapy lines. Seven had received prior anti-CD19 and/or anti-CD22 therapies, three had relapsed following approved CAR19 T-cell products, and six had experienced relapse after alloHSCT. The number of prior relapses ranged from 0 to 6, with a median interval of 51.5 months (range, 8-72 months) from initial diagnosis to CART22.19 infusion. The disease status prior to lymphodepletion was heterogeneous: Six infusions were given for MRD-level disease, five for overt hematologic relapse, one for progressive extramedullary disease with bone marrow remission (P1), and one for isolated CNS involvement without bone marrow infiltration (P5).

**Table 1:**
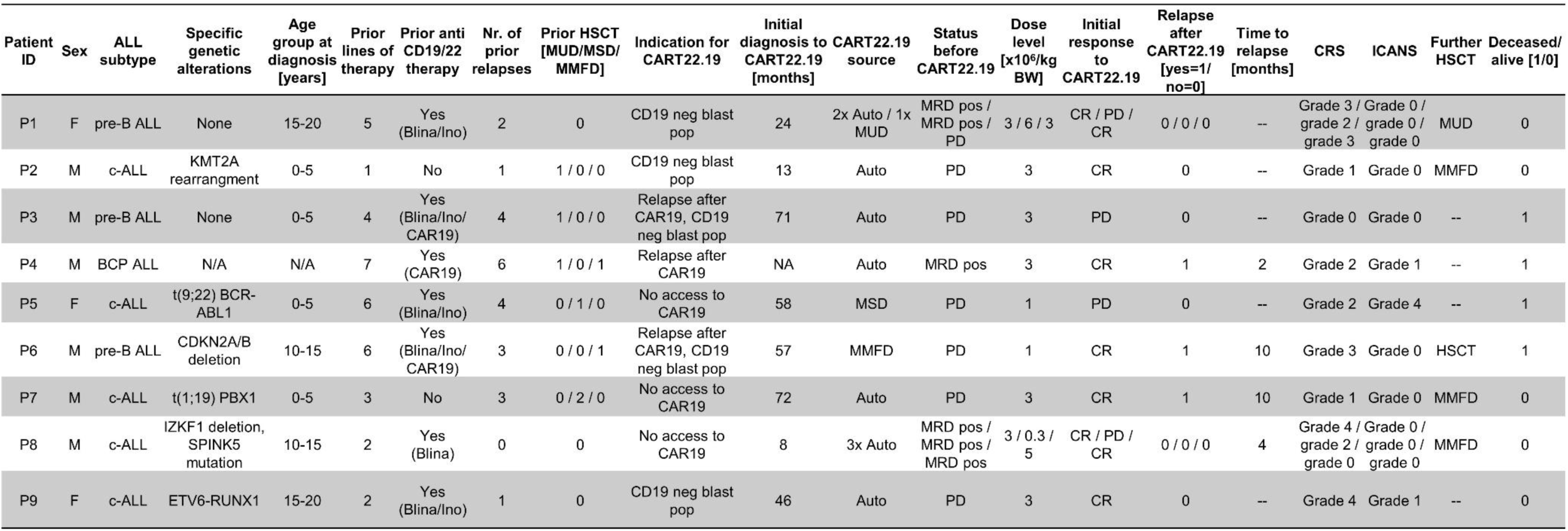
Clinical Characteristics and Outcomes of Pediatric Patients Treated with CART22.19. Allo: allogeneic; ALL: acute lymphoblastic leukemia; Auto: autologous; BCP: B-cell precursor; Blina: blinatumomab; BW: body weight; c-ALL: common acute lymphoblastic leukemia; CAR19: CD19 directed CAR T-cells; CR: complete remission; CRS: cytokine release syndrome; F: female; HSCT: hematopoietic stem cell transplantation; ICANS: immune effector cell-associated neurotoxicity syndrome; Ino: inotuzumab ozogamicin; M: male; MMFD: mismatched family donor; MRD: minimal residual disease; MSD: matched sibling donor; MUD: matched unrelated donor; neg: negative; NR: no remission; PD: persistent/progressive disease; pop: population; pos: positive; pre-B ALL: precursor B acute lymphoblastic leukemia.

Collectively, this cohort represents a highly heterogeneous and heavily pretreated group of non-selected pediatric patients with no remaining curative treatment options. The named-patient use of CART22.19 in this context highlights a pressing unmet clinical need.

### Adverse Events

The safety profile of CART22.19 therapy was evaluated across all 13 infusions (**Table 2**). Treatment-emergent toxicities within the first 28 days post-infusion were graded according to CTCAE and ASTCT criteria (27, 28). Adverse events (AEs) of any grade occurred following 11 of 13 infusions, most frequently pyrexia (84.6%), alanine aminotransferase (ALT) elevation (69.2%), and aspartate aminotransferase (AST) elevation (61.5%). Grade ≥3 AEs were observed after 10 infusions, most commonly neutropenia (76.9%), anemia (69.2%) and hypotension (38.5%). All grade 3 cytopenia resolved within 2 months. Grade ≥3 CRS occurred in five cases (38.5%) and resolved within two weeks following standard management per institutional guidelines. ICANS was observed in two patients, both presenting with mild symptoms (headache, dizziness). One patient (P5) with pre-existing CNS leukemia experienced recurrent seizures and somnolence before and after CART22.19 infusion. In the absence of CRS, elevated cytokines, or other ICANS features, these events were attributed to disease progression. CRS and ICANS were managed with tocilizumab (77%), dexamethasone (46%), and/or anakinra (15%), with complete resolution in all cases. Additional isolated events included sepsis, pancreatitis, and transient adrenal insufficiency. No cases of treatment-related death or HLH were reported. Overall, CART22.19 therapy demonstrated a manageable safety profile, with adverse events effectively controlled using established supportive care protocols.

**Table 2:** Adverse Events. AEs were graded per CTCAE v5.0 (28); CRS and ICANS per ASTCT criteria (27). Abbreviations: ALT: alanine aminotransferase; AST: aspartate aminotransferase; CRS: cytokine release syndrome; ICANS: immune effector cell-associated neurotoxicity syndrome; LDH: lactate dehydrogenase.

| | Any Grade | Grade $\geq 3$ |
| --- | --- | --- |
| **All adverse events** | 13 (100%) | 13 (100%) |
| Neutropenia | 2 (15.4%) | 10 (76.9%) |
| Anaemia | 3 (23.1%) | 9 (69.2%) |
| Pyrexia | 11 (84.6%) | 0 (0%) |
| ALT increase | 9 (69.2%) | 0 (0%) |
| AST increase | 8 (61.5%) | 0 (0%) |
| Hypoalbuminemia | 7 (53.8%) | 0 (0%) |
| LDH increase | 6 (46.1%) | 0 (0%) |
| Hypotension | 0 (0%) | 5 (38.5%) |
| Myalgia | 0 (0%) | 3 (23.1%) |
| Hypoxia | 2 (15.4%) | 1 (7.7%) |
| Necessity of<br>parenteral nutrition | 3 (23.1%) | 0 (0%) |
| Hypofibrinogenemia | 2 (15.4%) | 0 (0%) |
| Creatinine increase | 2 (15.4%) | 0 (0%) |
| Delir | 1 (7.7%) | 2 (15.4%) |
| Mucositis | 0 (0%) | 2 (15.4%) |
| Alkaline phosphatase<br>increase | 2 (15.4%) | 0 (0%) |
| Diarrhea | 2 (15.4%) | 0 (0%) |
| Headache | 2 (15.4%) | 0 (0%) |
| Neuropathic pain | 2 (15.4%) | 0 (0%) |
| Seizure | 0 (0%) | 1 (7.7%) |
| Sepsis | 0 (0%) | 1 (7.7%) |
| Pancreatitis | 1 (7.7%) | 0 (0%) |
| Transient adrenal<br>insufficiency | 1 (7.7%) | 0 (0%) |
| **CRS** | 6 (46.1%) | 5 (38.5%) |
| **ICANS** | 2 (15.4%) | 0 (0%) |
| **Death** | 0 (0%) | 0 (0%) |

### Clinical Outcomes

The clinical courses of the nine pediatric patients treated with CART22.19 are summarized in **Figure 2A**, illustrating treatment responses, remission durability, and overall outcomes in this high-risk, heavily pretreated cohort. As of the data cut-off (March 31, 2025), median follow-up was 23.2 months (range, 9.4-64.4). At the initial response assessment (30-60 days post-infusion), seven of nine patients had achieved complete molecular remission (**Figure 2B**). Three patients (P1, P2, P9) remained in ongoing remission. Notably, P1 achieved CR after receiving allogeneic CART22.19 following relapse and non-response to two previous autologous infusions and alloHSCT in non-remission. P2 achieved complete remission after a single CART22.19 infusion and subsequently underwent consolidative HSCT. P9 remains in remission without additional therapy. Four patients achieved MRD-negative CR but relapsed subsequently. Among responders, the median remission duration was 8.2 months (range: 1.3-63.3 months), with a median time to relapse of 3.6 months (range: 1.3-9.2 months). Two of these patients (P6, P7) underwent alloHSCT after relapse, and P7 remained in remission at last follow-up. Patient P8 achieved MRD-negative CR following two autologous CART22.19 infusions but experienced two relapses, indicating only transient disease control. Two patients (P3 and P5) did not achieve CR following CART22.19 therapy and succumbed to disease progression shortly after treatment. Notably, all patients with prior refractoriness to CAR19 T-cell therapy relapsed within 10 months of CART22.19 infusion. At the time of data cut-off, five of the nine patients (55.5%) were alive.

**Figure 2:**
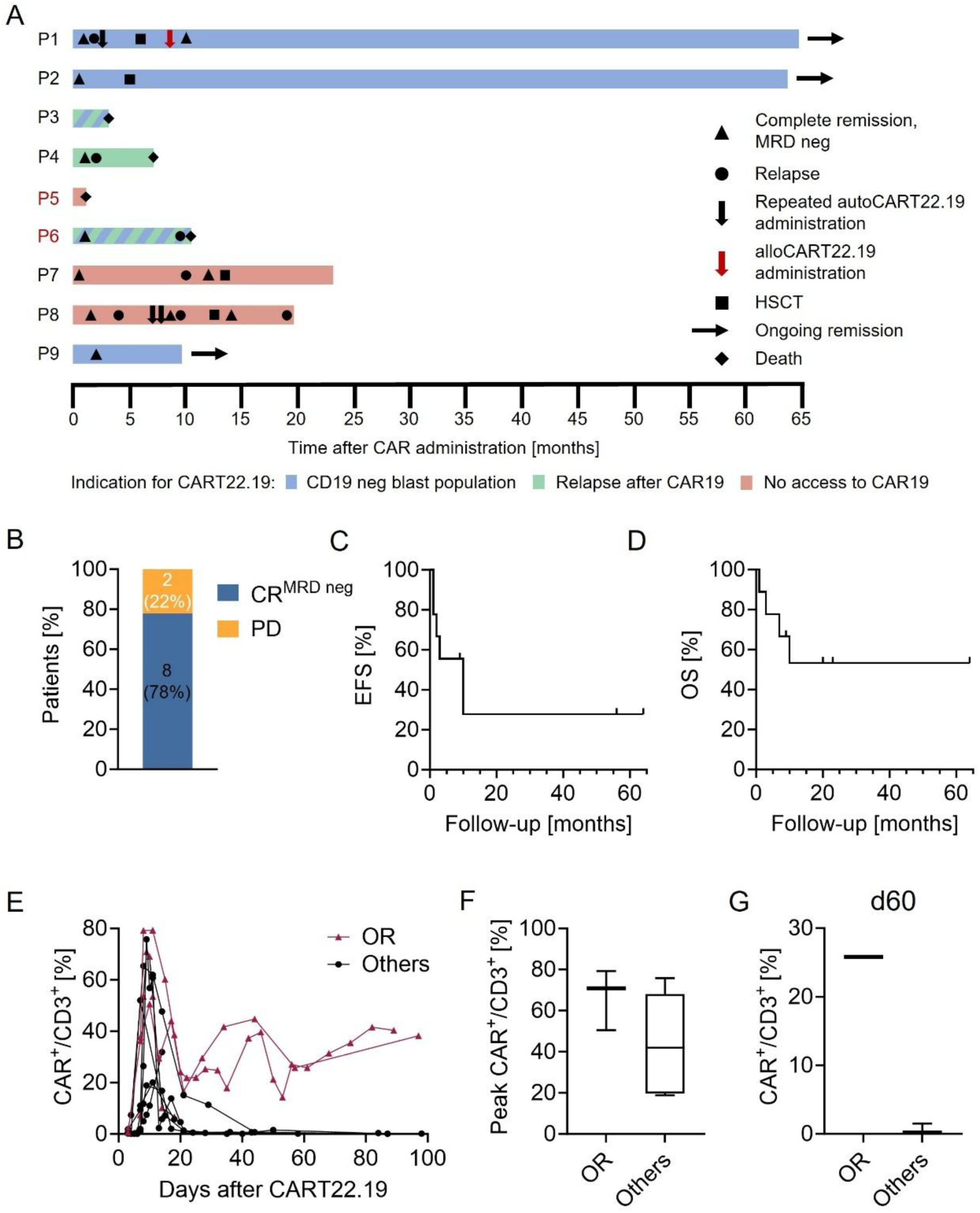
Clinical Outcomes following CART22.19 Therapy. **A** Swimmer plot illustrating the post-infusion clinical course for each patient. The indication for CART22.19 therapy is color-coded: blue indicates a CD19 negative blast population, green relapse after prior CAR19 therapy, and red cases where patients had no access to CAR19 therapy. Patient numbers in red refer to allogeneic CART22.19 recipients; black numbers indicate autologous CART22.19 recipients. **B** Initial response rates following CART22.19 administration, categorized as complete remission or persistent/ progressive disease. For patients who received multiple CART22.19 infusions, the response is assigned to the most recent infusion. **C** EFS after CART22.19 administration within the observation period; events were defined as molecular or clinical relapse or death. **D** OS of patients following CART22.19 therapy. **E** Longitudinal percentage of CAR⁺ cells within the CD3⁺ compartment after CART22.19 administration; patients with ongoing remission are shown in red, others in black. **F** Peak percentage of CAR^+^ cells within CD3^+^ T-cells, comparing patients with ongoing remission to others. **G** Frequency of CAR^+^ cells within CD3^+^ T-cells on day 60 post-infusion, comparing ongoing patients with remission to others. EFS and OS (panels C, D) were estimated using the Kaplan-Meier method; comparisons in panels F and G showed no significant differences (Mann-Whitney U test). Abbreviations: CAR: chimeric antigen receptor; CAR19: CD19-directed CAR; CART22.19: fully human tandem CAR T-cells with anti-CD22 and anti-CD19 scFvs; CR: complete remission; d60: day 60 post-infusion; EFS: event-free survival; HSCT: hematopoietic stem cell transplantation; neg: negative; OR: Ongoing remission; OS: overall survival; PD: persistent/progressive disease.

Kaplan-Meier analysis showed EFS rates of 55.6% at 6 months (95% CI, 20.4%-80.5%) and 27.8% at both 12 and 24 months (95% CI, 4.4%-59.1%), with events defined as molecular or clinical relapse or death (**Figure 2C**). OS rates were 77.8% at 6 months (95% CI, 36.5%-94%) and 53.3% at both 12 and 24 months (95% CI, 17.7%-79.6%) (**Figure 2D**). Median OS was not reached. Taken together, these results demonstrate that tandem CART22.19 therapy can induce complete and durable remissions in a subset of heavily pretreated pediatric B-ALL patients.

### Immunophenotypic Characteristics and CART22.19 Kinetics

Immunophenotypic analysis of leukemic blasts prior to CART22.19 infusion revealed heterogeneous target antigen expression. CD19 expression was positive in three patients (P4, P7, P8), heterogeneous in three (P1, P2, P6), and absent in two patients (P3 and P9) with isolated CD22⁺CD19⁻ disease (**Suppl. Figure S2**). CD22 expression was positive in six patients and heterogeneous in one (P1). No antigen expression data prior to treatment were available for P5, and CD22 expression was not assessed in P6.

Among the five patients who relapsed, one developed CD19⁻CD22⁺ disease after initially co-expressing both targets, while two relapsed with persistent CD19⁺CD22⁺ expression. Antigen expression data at relapse were not available for the remaining two patients. Notably, P9 achieved an ongoing remission despite complete absence of CD19 expression, highlighting the therapeutic contribution of the CD22-targeting component of the CAR22.19 construct.

CART22.19 expansion in peripheral blood was observed following 11 of 13 infusions, with peak levels occurring between days 7 and 15 (median, day 9). Peak frequencies of CART22.19 among peripheral T-cells ranged from 18.9% to 79.3%. Two patients with ongoing response maintained CART22.19 frequencies of over 40% at day 90 post-infusion. No data beyond day 20 were available for the third patient in ongoing remission (**Figure 2E**). In contrast, patients who did not achieve long-term remission exhibited low or rapidly declining CART22.19 frequencies. The mean peak CART22.19 frequency was 66.9% in ongoing responders compared to 44% in others (**Figure 2F)**, and by day 60, mean frequencies were 25.8% versus 0.63%, respectively (**Figure 2G**). Although these findings suggest a potential association between CART22.19 persistence and sustained clinical response, statistical significance was not reached due to limited sample size.

In summary, sustained remission in a patient with CD19-negative disease highlights the therapeutic relevance of CD22 targeting, while the association between declining CAR T-cell levels and relapse implicates limited CART22.19 persistence as a principal mechanism of treatment failure.

### Exploratory Analysis of Factors Associated with Ongoing Remission

An exploratory subgroup analysis was performed to identify clinical factors potentially associated with ongoing remission following CART22.19 therapy (**Suppl. Figure S3**). Among the nine pediatric patients treated, three (33%) achieved ongoing remission at a median follow-up of 23.2 months.

All continued responders were from the subgroup with CD19-negative leukemic subpopulations (60% [95% CI, 14.7-94.7%]), whereas no patient treated for relapse after CAR19 T-cell therapy or for lack of access to commercial products achieved an ongoing response (0% in both groups [95% CI, 0-56.1%]). Sustained remission was also more frequent in patients with fewer than three prior relapses (50% [95% CI, 15–85]) and in those treated within 50 months of initial diagnosis (75% [95% CI, 30.1-95.4]) compared with later treatment (0% [95% CI, 0-49.0]).

Other baseline characteristics, including age, immunophenotype (common vs. pre-B ALL), and number of prior treatment lines, showed no apparent association with treatment outcome. Given the limited sample size, none of these trends reached statistical significance and should be interpreted with caution.

Nevertheless, these findings suggest that CD19-negative leukemic subpopulations, shorter intervals from initial diagnosis to CART22.19 treatment, and fewer prior relapses may be associated with improved long-term outcomes. These observations warrant confirmation in larger patient cohorts.

### Long-term Remission after Combined RIC AlloHSCT and Donor-derived CART22.19

A detailed case presentation of P1 is provided, illustrating an innovative strategy to overcome resistance to CART22.19 (**Figure 3A**). When enrolled in the named-patient program, P1 presented with refractory pre-B ALL, including extensive skeletal involvement detected by ¹⁸F-FDG-PET-MRI (**Figure 3B; Suppl. Fig. S4A**) and CD19-negative blast populations, after five prior lines of therapy including inotuzumab ozogamicin and blinatumomab. Following standard lymphodepletion with fludarabine and cyclophosphamide (Flu/Cy), fresh autologous CART22.19 cells (autoCART22.19) were infused at a dose of 3 × 10⁶ CAR⁺ cells/kg BW. Grade 3 CRS developed and was effectively managed with tocilizumab and transient vasopressor support, resolving within two weeks. No neurotoxicity was observed. The patient achieved radiologic and molecular complete remission with a peak CAR T-cell expansion of 60% of viable T-cells (996 CAR T-cells/μL) on day 9 and detectable levels persisting through day 77 (**Figure 3C; Suppl. Fig. S4C**). Relapse occurred three months after autoCART22.19. A second infusion of cryopreserved autoCART22.19 did not show signs of clinical efficacy, no CRS, no CART22.19 expansion and no objective response. Re-staging revealed progressive disease with bulky lymphadenopathy in multiple regions (**Figure 3B**). Despite active disease, alloHSCT was performed using a TCRαβ/CD19-depleted graft from a 9/10 HLA-matched unrelated donor following bridging chemotherapy and RIC, without post-transplant immunosuppression.

**Figure 3:**
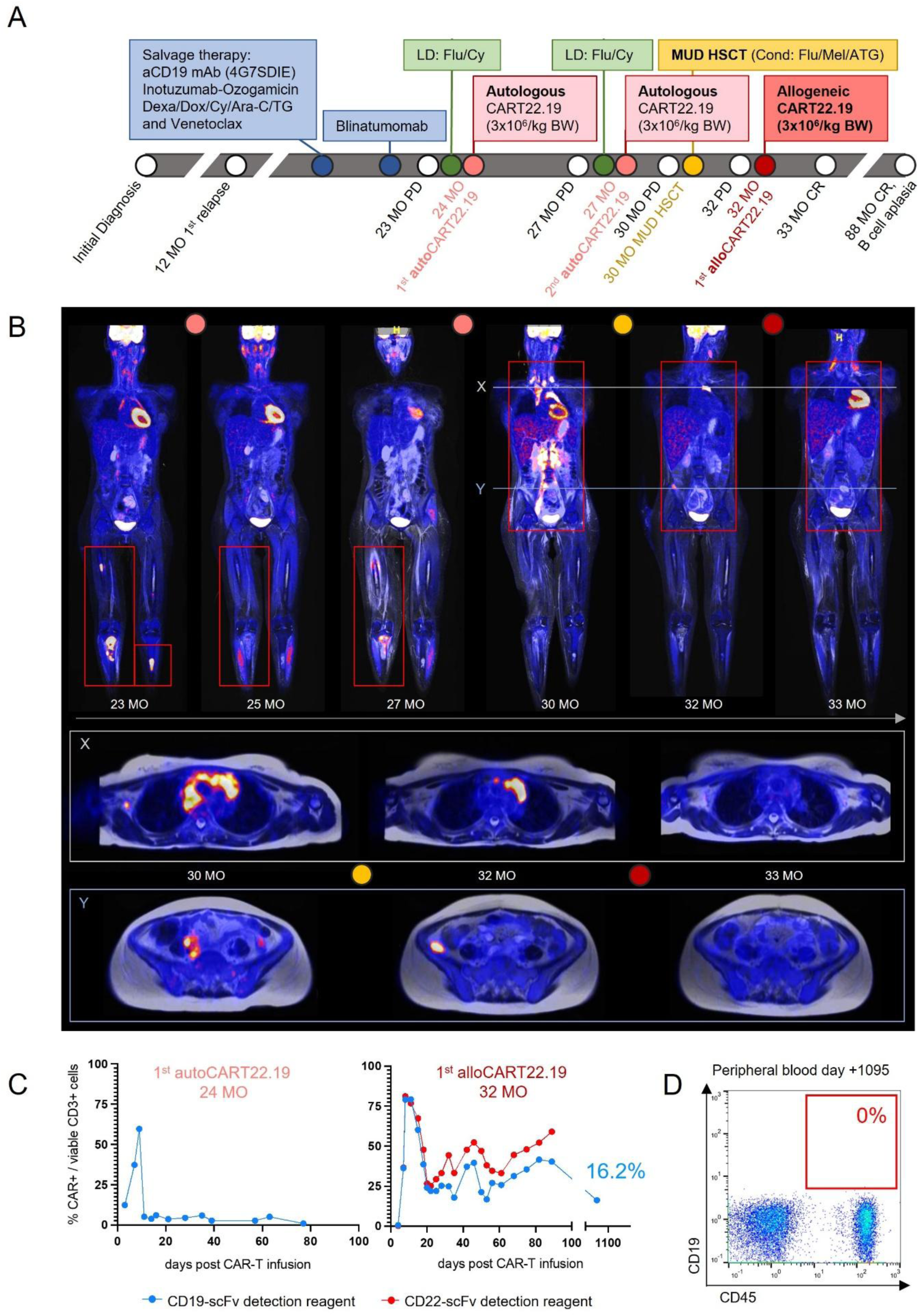
Allogeneic CART22.19 after RIC alloHSCT: Clinical Outcomes and CAR T Kinetics. **A** Schematic overview of the clinical course of patient P1. Time points indicate months since initial diagnosis. **B** Whole-body ^18^F-FDG PET-MRI at selected time points, referenced to months since initial diagnosis. The upper panel shows coronal images with red boxes marking sites of disease manifestation. X and Y indicate the axial slices shown in the middle and lower panels. Colored dots correspond to the timing of therapeutic interventions shown in panel A. **C** Longitudinal monitoring of CART22.19 in peripheral blood. Flow cytometry of PBMCs at the indicated time points post-infusion, with CAR⁺ cells shown as a percentage of CD3⁺ T-cells using CD19-(blue) and CD22-scFv–specific reagents (red). The left panel depicts expansion after the first autologous CART22.19 infusion, and the right panel after the allogeneic CART22.19 infusion. **D** Flow cytometric analysis of peripheral blood on day +1095 post-infusion, demonstrating persistent B-cell aplasia. CD19 is plotted against CD45. Abbreviations: ^18^F-FDG: fluorine-18 fluorodeoxyglucose; alloHSCT: allogeneic hematopoietic stem cell transplantation; ATG: antithymocyte globulin; Ara-C: cytarabine; BW: body weight; CAR: chimeric antigen receptor; CART22.19: fully human tandem CAR T-cells with anti-CD22 and anti-CD19 scFvs; CR: complete remission; Cy: cyclophosphamide; Dexa: dexamethasone; Dox: doxorubicin; Flu: fludarabine; LD: lymphodepletion; mAb: monoclonal antibody; Mel: melphalan; MO: months; MUD: matched unrelated donor; PD: progressive disease; PET-MRI: positron emission tomography-magnetic resonance imaging; RIC: reduced-intensity conditioning; scFv: single-chain variable fragment; TG: thioguanine.

Post-transplant evaluation demonstrated persistent extramedullary disease and rising MRD (2×10⁻⁴). On day +72 post HSCT, the patient received fresh donor-derived allogeneic CART22.19 (alloCART22.19) at a dose of 3 × 10⁶ CAR⁺ cells/kg BW, without additional lymphodepletion. Grade 3 CRS occurred but was effectively controlled and no ICANS or GvHD exacerbation was observed (**Suppl. Fig. S4B**). By day +17 post CART22.19, complete molecular and radiologic remission was achieved (**Figure 3B**).

Immune monitoring demonstrated distinct CAR T-cell expansion kinetics across the three infusions. The alloCART22.19 product peaked at 67% (1,110 CAR T-cells/μL) on day +15 and remained at 851 cells/μL through day +84 (**Figure 3C and Suppl. Fig. S2C**). At 4.5 years post-alloCART22.19 therapy, the patient remains in ongoing complete remission. AlloCART22.19 continue to comprise 16% of circulating T-cells and retain functional activity, as evidenced by sustained B-cell aplasia (**Figure 3D**). This case illustrates the potential of combined RIC HSCT and donor-derived CART22.19 therapy to induce long-term remission in refractory disease.

## Discussion

This report presents the preclinical validation and initial clinical experience with CART22.19, a fully human tandem CAR T-cell therapy targeting CD19 and CD22, in a cohort of heavily pretreated pediatric patients with relapsed or refractory B-ALL. *In vitro* and *in vivo* analyses confirmed potent, antigen-specific cytotoxicity and dual-target activation. Clinically, CART22.19 was administered in a named-patient program to nine pediatric patients lacking standard curative treatment options.

The safety profile of CART22.19 was manageable and consistent with other published CAR T-cell studies in pediatric B-ALL including the ELIANA study, which led to the approval of tisagenlecleucel (6, 17–21, 24). Based on ASTCT criteria, grade ≥3 CRS occurred in 38.5% of infusions, a rate that remained consistent when retrospectively applying the University of Pennsylvania grading scale used in ELIANA (29). While ELIANA reported grade 3-4 CRS in 25% and grade ≥3 neurotoxicity in 13.3% of patients, no severe ICANS occured in our cohort (6). Notably, no treatment-related deaths were observed, in contrast to a study by Wang et al., which reported three fatalities among 225 patients receiving coadministered CD19- and CD22-directed CAR T-cells (17). These findings suggest a favorable safety profile of CART22.19 consistent with both single- and dual-antigen approaches.

The clinical outcomes observed in our CART22.19 cohort align with those reported in other dual-targeting CAR T-cell trials (18, 19, 21). Initial molecular complete remission was achieved in 78% of patients, with 12-month EFS and OS rates of 27.8% and 53.3%, respectively. These outcomes are consistent with the bicistronic CD19/CD22 CAR trial by Cordoba et al. (67% CR, 30% EFS at 12 months). Initial CR rates were also comparable to those reported in the ELIANA study (81%), despite our cohort having received more prior lines of therapy and including patients with CD19-negative disease and/or refractory to tisagenlecleucel. Taken together, these findings highlight the clinical potential of CART22.19 in high-risk pediatric B-ALL populations with otherwise limited treatment options.

Several dual-targeting CAR T-cell strategies engaging CD19 and CD22 have shown limited CD22-mediated functionality, with reduced cytokine release and cytotoxicity upon CD22 engagement in bicistronic models, and a predominant dependence on CD19 targeting in co-transduction and coadministration approaches (17–19). In contrast, our CART22.19 construct consistently elicited robust T-cell activation and cytokine secretion in response to both CD19⁺ or CD22⁺ targets *in vitro*. Moreover, the achievement of a complete and sustained molecular remission in a patient with CD19-negative disease provides compelling *in vivo* evidence of effective CD22-mediated antileukemic activity. These findings support the functional competence of the CD22-binding domain in our tandem CAR and highlight its potential to overcome limitations observed in previous approaches.

Despite an initial complete molecular remission rate of 78%, four patients relapsed. In one case, relapse occurred with CD19-negative disease. In contrast, two patients relapsed with CD19⁺CD22⁺ disease and low levels of CART22.19 detectable in peripheral blood, suggesting insufficient functional CAR T-cell persistence as the likely cause of relapse. This pattern is consistent with observations by Ghorashian et al. and others, in which most relapses occurred despite sustained target antigen expression (17–19, 21).

The mechanisms underlying reduced CAR persistence remain incompletely understood. Our CART22.19 product exhibited a central memory T-cell phenotype *in vivo* at day 28 post-infusion, a profile previously associated with favorable persistence. However, it may lack the long-term durability of stem-like memory or early central memory T-cell subsets (30–33). Additionally, tandem CAR constructs may be susceptible to functional limitations arising from tonic signaling (20, 21). In our cohort, limited post-infusion monitoring and immunophenotyping restricted comprehensive long-term functional assessment of CART22.19. Although immune-mediated rejection has been proposed in previous studies as a potential cause of limited persistence, this appears less likely, given the use of a fully humanized CAR construct (34, 35).

Notably, Das et al. recently demonstrated that T-cells collected after multiple cycles of chemotherapy exhibit mitochondrial dysfunction and impaired metabolic reserve, significantly limiting CAR T-cell expansion and persistence (36). These findings highlight that the intrinsic quality of autologous T-cells, particularly in heavily pretreated patients, may constrain the therapeutic efficacy of CAR T-cell approaches. This underscores the multifactorial nature of treatment failure and highlights the relevance of strategies aimed at enhancing T-cell fitness. One approach to improving efficacy involves the use of donor-derived CAR T-cell therapy, which leverages the proliferative and functional advantages of immunocompetent T-cells from healthy donors. The clinical course of patient P1 illustrates the therapeutic benefit of this strategy. After two unsuccessful autologous CART22.19 infusions, the patient received RIC alloHSCT in non-remission followed by donor-derived CART22.19 and subsequently achieved long-term remission. In contrast to other allogeneic strategies involving TCR editing to prevent graft-versus-host-disease (GvHD), our strategy preserved the native TCR, minimizing manipulation that may impair CAR T-cell function (37, 38). Importantly, no unexpected toxicities or GVHD occurred, and durable B-cell aplasia was achieved beyond three years after infusion. Consistent with our observations, prior studies in B-ALL and T-cell malignancies have demonstrated that donor-derived CAR T-cell products can induce deep and durable remissions, with superior expansion, persistence, and immune fitness compared to autologous products, particularly in heavily pretreated patients (39–42). Together, these data support the safety and efficacy of combining RIC alloHSCT with donor-derived CAR T-cells and establish a promising platform for delivering potent, long-lasting cellular therapies for highly pretreated patients.

Despite these encouraging findings, several limitations must be acknowledged. The named-patient framework introduced heterogeneity in patient characteristics, disease burden, prior therapies and antigen expression. Case-by-case decision-making, driven by clinical urgency, limited consistency in treatment and follow-up. The small cohort size and incomplete post-relapse immunophenotyping reduced statistical power and mechanistic insight. Consequently, these results should be interpreted with caution and require validation in prospective, controlled clinical trials.

### Conclusion

CART22.19 demonstrated a favorable safety profile and promising clinical activity in high-risk pediatric B-ALL. Durable remissions were achieved in a subset of patients, even in the absence of CD19 expression, while relapse despite preserved antigen expression highlights limited CAR T-cell persistence as a key challenge. Our findings support the feasibility and therapeutic potential of donor-derived CAR T-cells following RIC alloHSCT to address the limitations of autologous products. Further clinical evaluation of CART22.19 is warranted, with particular emphasis on allogeneic approaches to enhance persistence and improve outcomes in heavily pretreated pediatric leukemia.

## Data Availability

All data produced in the present study are available upon reasonable request to the authors

## Acknowledgements

The authors sincerely thank the patients and their families for their trust and participation in this named-patient use program. We also thank the referring physicians and the clinical staff of the University Children’s Hospital Tuebingen for their expertise in the clinical management of the patients. We acknowledge Miltenyi Biotec GmbH for providing GMP-grade CAR22.19 vector.

## Authorship Contributions

PL, PS, PH, DS, RH and CMS conceptualized and supervised the project. ASM and CMS wrote the manuscript. PH, DS and RO designed the CAR22.19, performed the pre-clinical experiments and contributed to data analysis and interpretation. ASM, PL, PS, FC, SS, FS, ME, MD, JS, JHS, RH and CMS, designed and performed the clinical study, response assessment and analyzed and/or interpreted the data. DA, SS, SK, CB, FS, MS, LH and BA generated the clinical CAR22.19 products and performed quality controls and immune monitoring. All authors contributed to the preparation of the manuscript and approved the submitted version.

