## Supplementary Data for "Human CART22.19 Therapy in Refractory Pediatric B-ALL: Insights from a Named-Patient Cohort"

### Supplementary Methods

##### Design, Vector Production and Preclinical Generation of CAR22.19

For comparative analyses, a second-generation CAR19 construct was generated alongside the tandem CAR22.19 construct. Both vectors shared an identical lentiviral backbone, with the CAR19 construct containing only the anti-CD19 scFv. CAR sequences were synthesized as gBlocks (Integrated DNA Technologies, Coralville, IA, USA) and cloned into a third-generation lentiviral vector plasmid (Lentigen Technology, Gaithersburg, MD, USA) regulated by the human elongation factor 1α (EF-1α) promoter. For the preclinical evaluation, lentiviral particles were produced by transient transfection of HEK 293T cells using a four-plasmid packaging system, as previously described (1). Viral supernatants were harvested, concentrated by centrifugation, and stored at -80 °C until use.

T-cells were isolated from leukapheresis products (HemaCare, Northridge, CA) or buffy coats (Oklahoma City, OK, USA). Human primary $CD4^+$ or $CD8^+$ cells were isolated from blood products by immunomagnetic separation according to manufacturer's protocol (Miltenyi Biotec, Bergisch Gladbach, Germany), cryopreserved, and later thawed for use. Upon thawing, T-cells were activated using CD3/CD28 MACS® GMP T-Cell TransAct reagent and cultured in TexMACS GMP medium (Miltenyi Biotec) supplemented with 30 IU/mL IL-2 at a density of $1 \times 10^6$ cells/mL. Lentiviral transduction was performed one day after activation. On day 3, cells were washed and resuspended in fresh IL-2-supplemented TexMACS™ medium. Cell cultures were maintained at a density of 0.5-2 × $10^6$ cells/mL and harvested between days 8 and 10.

##### Flow Cytometric Analysis

Flow cytometry was used to assess surface molecule expression on CAR T-cells. Cells were stained in cold AutoMACS buffer with 0.5% bovine serum albumin (Miltenyi Biotec), washed, and resuspended in 200 µL Running Buffer for acquisition on a MACSQuant®10 Analyzer. CAR expression was determined using CD22-his (Abcam) or CD19-Fc (R&D Systems) peptides, followed by secondary staining with anti-His-PE (Miltenyi Biotec) or anti-Fc-AF647 (Jackson ImmunoResearch). T-cell subsets were analyzed with anti-CD4 VioBlue (clone VIT4), and dead cells excluded using Viobility 405/520 Fixable Dye (Miltenyi Biotec).

**_In Vitro_ Cytotoxicity and Cytokine Secretion Assays**

Cytotoxicity assays were performed to evaluate the functional activity of CAR T-cell constructs. CAR T-cells were co-cultured for 16 hours with 5,000 target cells stably expressing firefly luciferase at the indicated effector to target (E:T) ratios. Following incubation, Steady-Glo® reagent (Promega, Madison, WI, USA) was added to each well, and luminescence was measured using a GloMax Luminometer (Promega) and reported as counts per second (CPS). Maximum and minimum signals were established using target-only wells treated with or without 1% Tween-20, respectively. Specific lysis was calculated using the formula: % specific lysis = 1 - (sample CPS - min CPS) / (max CPS - min CPS)

Supernatants collected from co-cultures were analyzed for cytokine release using ELISA kits (Thermo Fisher Scientific, Waltham, MA, USA) to quantify IL-2 and IFN-γ concentrations.

**NSG mice experiments**

All animal experiments were conducted at MI Bioresearch (now part of LabCorp; Ann Arbor, MI, USA) in accordance with protocols approved by the Institutional Animal Care and Use Committee (IACUC) and in full compliance with applicable laws, regulations, and National Institutes of Health (NIH) guidelines. NSG mice (NOD.Cg-Prkdc^scid Il2rg^tm1Wjl/SzJ) were intravenously injected with $1 \times 10^6$ NALM-6 cells, a model of B-cell precursor leukemia. Tumor burden was monitored via bioluminescent imaging (BLI) using the Xenogen IVIS-200 system (PerkinElmer, Shelton, CT). On day 3 post-injection, mice with comparable tumor loads were randomized into treatment groups and received $1 \times 10^7$ CAR T-cells per mouse. Tumor progression was monitored at defined intervals (days 0, 3, 7, 14, 21, 28, and 37) using *in vivo* imaging. Peripheral blood was collected on day 28 after CAR T-cell administration and analyzed by flow cytometry.

**Quality Control of CART22.19 Products**

Before infusion, clincial CART22.19 products underwent standard quality control evaluations, including sterility testing, assessment of cell viability, and measurement of transduction efficiency. Microbiological testing was conducted in accordance with GMP guidelines. Mean cell viability at the time of product release was 99.7% (range, 99.4-99.8%). Transduction efficiency was evaluated by flow cytometry using CD19-Fc and CD22-his detection reagents, with a mean transduction rate of 56.7% (range, 48.3-70.8%) at harvest. A summary of in-process controls and final product testing is provided in Supplementary Table S1.

**Immunophenotyping of Leukemic Blasts**

Bone marrow aspirate samples obtained at initial diagnosis and relapse were analyzed by flow cytometry at accredited laboratories of the University Hospital Frankfurt, following the protocol described by Karawajew et al (2). Leukemic blasts were identified based on the expression profile of CD19, CD22, CD24, CD25, CD10, CD20, CD34, CD38, and CD44. Antigen expression intensity was evaluated in accordance with the guidelines established by Dworzak et al (3).

**Immune Monitoring**

Immune monitoring of CART22.19 was performed by flow cytometric analysis of peripheral blood samples collected at multiple time points following infusion. Red blood cells were lysed prior to staining. Subsequently, 100 µL of lysed whole blood was incubated with 2 µL of either CD19-CAR or CD22-CAR detection reagent (Miltenyi Biotec, Cat. #130-115-965 and #130-126-727) for 10 minutes at room temperature. Following two washes with CliniMACS PBS/EDTA buffer (Miltenyi Biotec), cells were stained with a fluorochrome-conjugated antibody panel consisting of anti-Biotin-PE (#130-110-951), CD45-VioBlue (#130-110-637), CD3-FITC (#130-113-138), CD4-VioGreen (#130-113-221), CD8-APC-Vio770 (#130-110-681), and 7-AAD viability dye (#130-111-568) for an additional 10 minutes.

Samples were acquired on a MACSQuant® flow cytometer (Miltenyi Biotec), and data were analyzed using FlowJo software version 10.8 (BD, Ashland, OR, USA). The gating strategy is provided in Supplementary Figure S4D.

**Therapy Response Assessment**

Therapeutic response was evaluated through serial bone marrow aspirates and, in patients with a history of CNS involvement, cerebrospinal fluid analysis. MRD in the bone marrow was assessed by real-time quantitative PCR (RQ-PCR) when patient-specific molecular markers were available, or by multiparameter flow cytometry otherwise. MRD evaluations were performed at accredited laboratories at University Hospital Frankfurt, with flow cytometric analysis conducted according to the protocol described by Karawajew et al (2). MRD negativity was defined as < $10^{-4}$ leukemic cells by PCR or < 0.01% blasts by flow cytometry. In cases of suspected or confirmed extramedullary involvement, imaging studies (including MRI, CT, PET-MRI, or PET-CT) were performed every three months or as clinically indicated. Complete remission was defined as the absence of detectable extramedullary disease and MRD negativity in the bone marrow.

CART22.19 persistence in peripheral blood was assessed by flow cytometry at variable intervals, as described in the Immune Monitoring section. Detection of CAR T-cells was defined as ≥ 0.01% of $CD3^+$ cells. CART22.19 measurements were performed for a median duration of 70 days (range: 14-176 days) following infusion. In parallel, peripheral blood flow cytometry for CD45, CD3, and CD19 was used to quantify circulating B-cells as a surrogate marker of CAR T-cell activity. Longitudinal kinetics of CAR T-cell persistence are shown in Figure 2.

**Safety Monitoring and Adverse Event Assessment**

CRS and ICANS were graded according to the 2019 consensus criteria of the ASTCT (4). All other AEs and clinically significant laboratory abnormalities were evaluated using the CTCAE version 5.0 (5).

Laboratory safety monitoring was conducted on blood samples obtained prior to, during, and following CART22.19 infusion. Routine assessments included blood counts, coagulation profiles, renal and hepatic function panels and inflammatory markers such as IL-2 and IL-6, measured every other day for a minimum of one month. Additional laboratory evaluations, including microbiological and infectious disease testing, were performed twice weekly or as clinically indicated throughout the post-infusion period.

Management of grade ≥3 CRS included intravenous dexamthasone, tocilizumab and/or anakinra. As seizure prophylaxis, some patients were preemptively treated with levetiracetam. Anemia and thrombocytopenia were managed with red blood cell and platelet transfusions, respectively.

### Supplementary Figures

#### Supplementary Figure S1

**A-B** CART22.19, CAR19, or UTD were co-cultured with target cell lines at indicated E:T ratios for 16 hours. Supernatants were collected and analyzed by ELISA to quantify IL-2 (**A**) and IFN-γ (**B**) secretion. Target cell lines included Raji WT (CD19$^{+}$CD22$^{+}$), Raji CD19$^{KO}$ (CD19$^{-}$CD22$^{+}$), Raji CD22$^{KO}$ (CD19$^{+}$CD22$^{-}$), and Raji CD19$^{KO}$CD22$^{KO}$ (CD19$^{-}$CD22$^{-}$). Data represent mean ± SD of three technical replicates. Statistical significance was determined using unpaired t-tests. **C-D** NSG mice were injected intravenously with $1\times10^{6}$ NALM-6 cells expressing firefly-luciferase on day 0, followed by intravenous infusion of $1\times10^{7}$ effector cells (CART22.19, CAR19, or UTD) on day 3. On day 28, peripheral blood was collected for flow cytometric analysis. **C** Total number of CAR$^{+}$ T-cells in 50 µL of peripheral blood. Each data point represents an individual mouse (n = 4-6 per group); data shown as mean ± SEM. **D** Total number CAR$^{+}$ central memory T-cells (CD62L$^{+}$CD45RO$^{+}$) in 50 µL of peripheral blood. Each data point represents an individual mouse (n = 4-6 per group); data shown as mean ± SEM. Statistical comparisons were performed using unpaired Student's t-tests. Abbreviations: CAR: chimeric antigen receptor; CAR19: CD19-directed CAR; CART22.19: fully human tandem CAR T-cells with anti-CD22 and anti-CD19 scFvs; Conc.: concentration; ELISA: enzyme-linked immunosorbent assay; E:T: effector-to-target ratio; IFN-γ: interferon gamma; IL-2: interleukin-2; KO: knockout; ns: not significant; NSG: NOD scid gamma; SD: standard deviation; SEM: standard error of the mean; UTD: untransduced activated T-cells; WT: wild type.

#### Supplementary Figure S2

Representative flow cytometry gating strategy exemplified for patient P9, as applied to all patients to assess CD19 and CD22 expression on leukemic blasts in bone marrow. **A** Leukocytes were first gated based on FSC-H vs. SSC-H, and blasts were identified as a CD45$^{dim}$ population expressing CD10, followed by evaluation of CD19 expression. **B** In a parallel analysis, leukocytes were gated as above, and blasts were defined as CD34$^{weak}$. Coexpression of CD19 and CD22 was then assessed. Sequential gating steps are indicated by red arrows. Abbreviations: FSC-H: forward scatter height; SSC-H: side scatter height.

**Supplementary Figure S3**

Subgroup analysis of patients with ongoing response following CART22.19 therapy. Forest plot illustrating the proportion of patients with ongoing remission (P1, P2, P9) across clinical and treatment-related subgroups. Stratification variables included age, disease classification, prior lines of therapy, number of previous relapses, history of allogeneic stem cell transplantation, indication for CAR T-cell therapy, interval from initial diagnosis to CAR T-cell infusion, and CAR T-cell dose level. Data are presented as percentages with 95% CI calculated using the Clopper-Pearson method. Each dot represents the mean proportion of patients with sustained response within the respective subgroup. Abbreviations: ALL: acute lymphoblastic leukemia; BW: body weight; CAR: chimeric antigen receptor; CART22.19: fully human tandem CAR T-cells with anti-CD22 and anti-CD19 scFvs; CI: confidence interval; HSCT: hematopoietic stem cell transplantation.

**Supplementary Figure S4**

Detailed clinical and immunological course of patient P1 is shown, including longitudinal monitoring of disease status, cytokine release, CAR T-cell dynamics, and representative flow cytometry gating strategy. **A** MRD levels in bone marrow, assessed by RT-PCR at the indicated time points, referenced to months since initial diagnosis. Colored dots correspond to the timing of therapeutic interventions summarized in Figure 3A. **B** Plasma cytokine levels post-infusion of CART22.19. Concentrations of IL-6 and sIL-2R were measured in peripheral blood plasma at the indicated time points. **C** Longitudinal monitoring of CART22.19 in peripheral blood. PBMCs were analyzed by flow cytometry post-infusion. The absolute number of viable CART22.19$^+$ cells per µL blood was determined using a CD19-scFv detection reagent. Data are shown for total T-cells (CD3$^+$, red), cytotoxic T-cells (CD3$^+$CD8$^+$, dark red), and T helper cells (CD3$^+$CD4$^+$, yellow). **D** Representative flow cytometry gating strategy used to assess CART22.19 in peripheral blood. Abbreviations: 7AAD: 7-aminoactinomycin D; CAR: chimeric antigen receptor; CART22.19: fully human tandem CAR T-cells targeting CD19 and CD22; FSC: forward scatter; IL-6: interleukin-6; MO: months; MRD: minimal residual disease; MUD: matched unrelated donor; PBMC: peripheral blood mononuclear cell; RT-PCR: reverse transcriptase polymerase chain reaction; scFv: single-chain variable fragment; sIL-2R: soluble interleukin-2 receptor; SSC: side scatter.

**Supplementary Table S1: Manufacturing Parameters and Product Characteristics of Patient-Specific CART22.19**

**Abbreviations**: transd: transduction.

**Supplementary Figure S1**

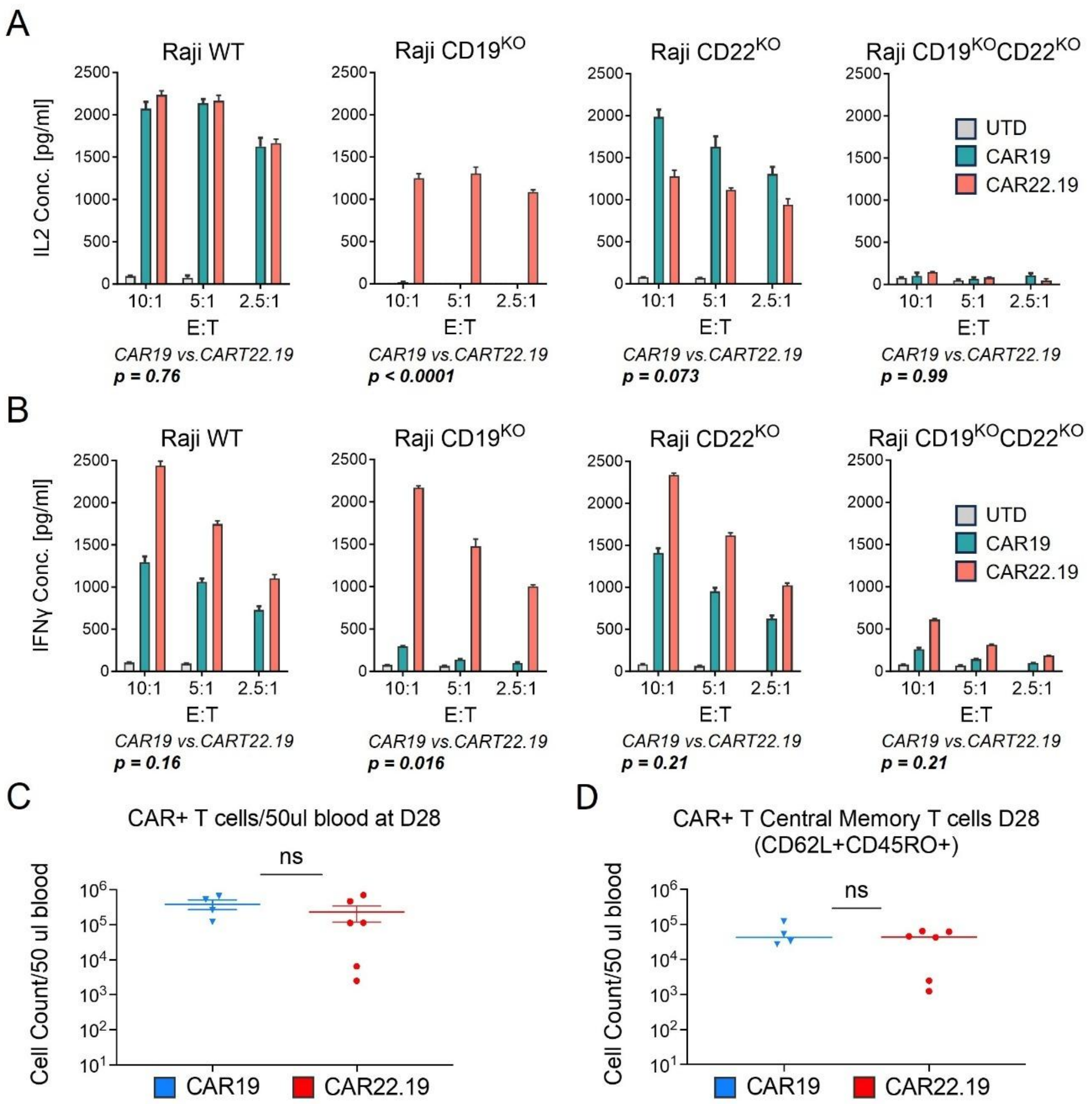

**Supplementary Figure S2**

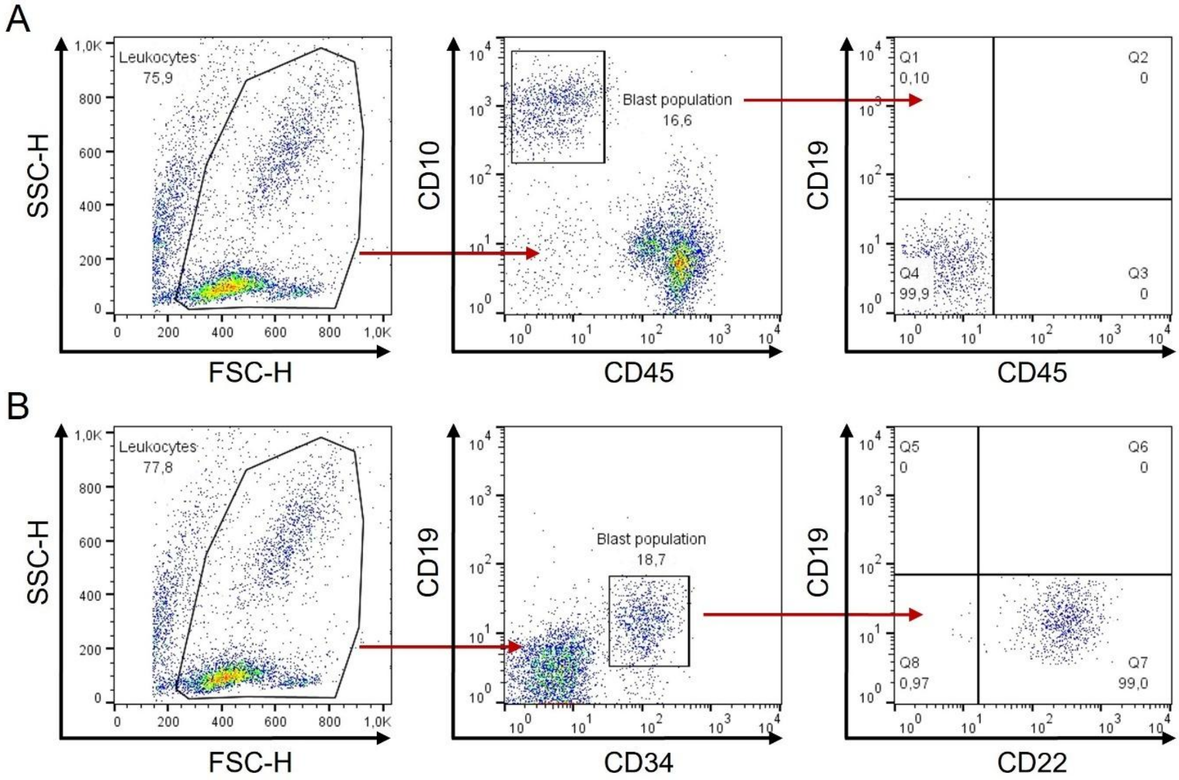

#### Supplementary Figure S3

| | Number of patients (n=9) | Number of patients in ongoing remission (n=3) | Percentage of patients in ongoing remission [95% CI] |
|---|---|---|---|
| **Age** | | | |
| < 10 years | 4 | 1 | 25 [4.6-69.9] |
| > 10 years | 4 | 2 | 50 [15-85] |
| **Disease classification** | | | |
| c-ALL | 5 | 2 | 40 [11.8-76.9] |
| pre-B ALL | 3 | 1 | 33.3 [6.1-79.2] |
| **Previous lines of therapy** | | | |
| ≤ 3 lines | 5 | 2 | 40 [11.8-76.9] |
| > 3 lines | 4 | 1 | 25 [4.6-69.9] |
| **Number of prior relapses** | | | |
| < 3 relapses | 4 | 2 | 50 [15-85] |
| ≥ 3 relapses | 5 | 1 | 20 [3.6-62.4] |
| **Relapse after allogenic HSCT** | 6 | 1 | 16.7 [3-56.4] |
| **Indication for CART22.19** | | | |
| Refractory to CAR19 | 3 | 0 | 0 [0-56.1] |
| CD19 negative blast population | 5 | 3 | 60 [14.7-94.7] |
| No access to CAR19 | 3 | 0 | 0 [0-56.1] |
| **Time from initial diagnosis to CART22.19** | | | |
| < 50 months | 4 | 3 | 75 [30.1-95.4] |
| > 50 months | 4 | 0 | 0 [0-49] |
| **Dose level** | | | |
| 3x $10^6$ $CAR^+$ cells/kg BW | 8 | 3 | 37.5 [13.7-69.4] |
| Other | 5 | 0 | 0 [0-43.4] |

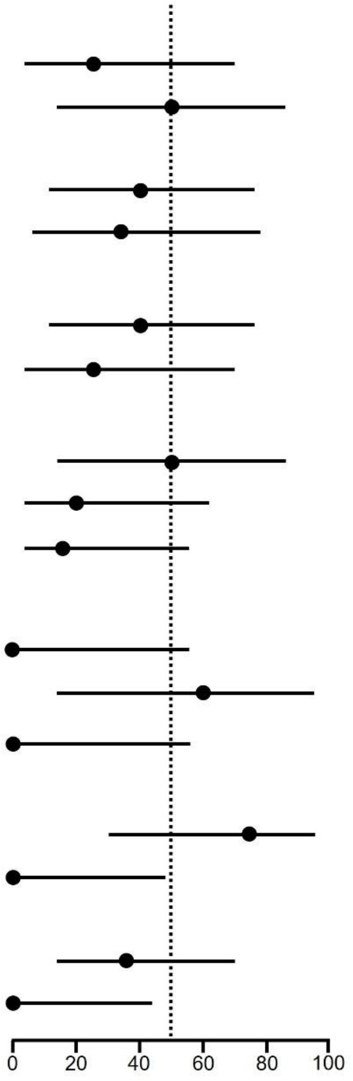

#### Supplementary Figure S4

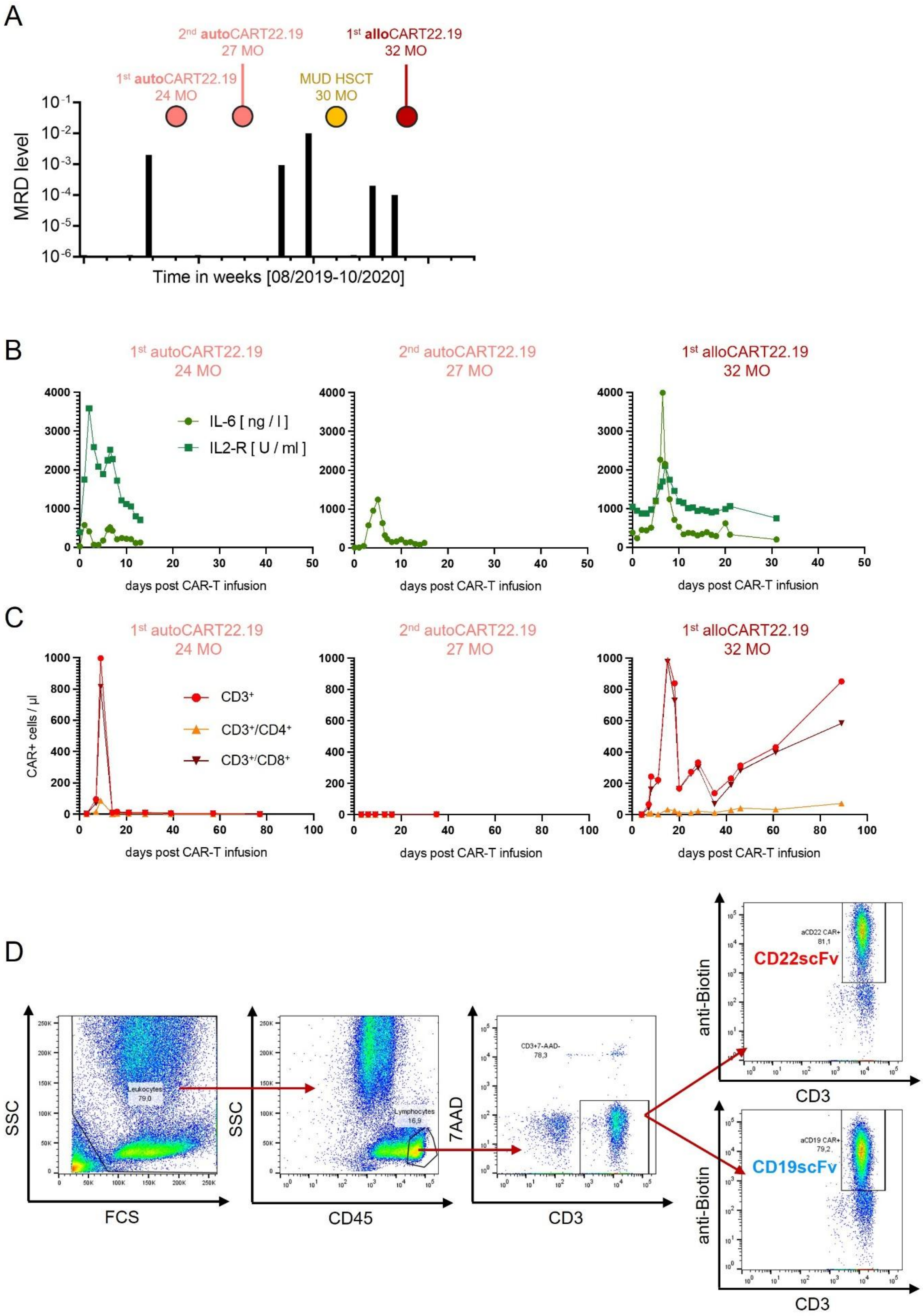

**Supplementary Table S1**

| Patient | Recovery after enrichment [%] | Day 5 cell count [$x10^6$] | Day 5 transd. efficiency | Harvest day | Harvest cell count [$x10^8$] | Harvest transd. frequency [%] | Vitality frequency [%] |
|---|---|---|---|---|---|---|---|
| P1 | 27.2 | 122 | 48.4 | 9 | 11.5 | 49 | 99.8 |
| P2 | 43.6 | 290 | 52 | 11 | 19.3 | 48.3 | 99.4 |
| P3 | 68.7 | 290 | 61.5 | 11 | 21.4 | 59.2 | 99.8 |
| P4 | 34 | 165 | 43.4 | 11 | 19.7 | 49.2 | 99.6 |
| P5 | 69.9 | 270 | 46.4 | 7 | 8.2 | 49.3 | 99.7 |
| P6 | 58.7 | 282 | 60.1 | 12 | 33.5 | 69 | 99.8 |
| P7 | 62.5 | 492 | 74 | 12 | 37.4 | 70.8 | 99.7 |
| P8 | 64.7 | 455 | 53 | 12 | 26.6 | 53.1 | 99.8 |
| P9 | 63.3 | 306 | 57.8 | 12 | 25.6 | 62 | 99.4 |
